# Clinical features, demography and predictors of outcomes of SARS-CoV-2 infection in a tertiary care hospital in India-A cohort study

**DOI:** 10.1101/2021.08.10.21261855

**Authors:** Arunmozhimaran Elavarasi, Hari Krishna Raju Sagiraju, Rohit Kumar Garg, Saurav Sekhar Paul, Brajesh Ratre, Prashant Sirohiya, Nishkarsh Gupta, Rakesh Garg, Anuja Pandit, Saurabh Vig, Ram Singh, Balbir Kumar, Ved Prakash Meena, Naveet Wig, Saurabh Mittal, Saurabh Pahuja, Karan Madan, Tanima Dwivedi, Nupur Das, Ritu Gupta, Ashima Jain Vidyarthi, Arghya Das, Rama Chaudhary, Laxmitej Wundawalli, Angel Rajan Singh, Sheetal Singh, Manisha Pandey, Abhinav Mishra, Karanvir Singh Matharoo, Sunil Kumar, Anant Mohan, Randeep Guleria, Sushma Bhatnagar

## Abstract

**Background:** The second wave of the COVID-19 pandemic hit India from early April 2021 to June 2021 and more than 400,000 cases per day were reported in the country. We describe the clinical features, demography, treatment trends, baseline laboratory parameters of a cohort of patients admitted at the All India Institute of Medical Sciences, New Delhi with SARS-CoV-2 infection and their association with the outcome.

**Methods:** This was a retrospective cohort study describing the clinical, laboratory and treatment patterns of consecutive patients admitted with SARS-CoV-2 infection. Multivariate logistic regression models were fitted to identify the clinical and biochemical predictors of developing hypoxia, deterioration during the hospital stay and death.

**Findings:** A total of 2080 patients were included in the study. The case fatality rate was 19.5%. Amongst the survivors, the median duration of hospital stay was 8 (5-11) days. Out of 853 (42.3%%) of patients who had COVID-19 Acute respiratory distress syndrome at presentation, 340 (39.9%) died. Patients aged 45-60 years [OR (95% CI): 1.8 (1.2-2.6)p =0.003] and those aged >60 years [OR (95%CI): 3.4 (2.3-5.2), p<0.001] had a higher odds of death as compared to the 18-44 age group. Vaccination reduced the odds of death by 30% [OR (95% CI): 0.7 (0.5-0.9), p=0.036]. Patients with hyper inflammation at baseline as suggested by leucocytosis [OR (95% CI): 2.1 (1.4-3.10), p <0.001], raised d-dimer >500 mg/dL [OR (95% CI): 3.2 (2.2-4.6), p <0.001] and raised C-reactive peptide >0.5 mg/L [OR (95% CI): 3.8 (1.1-13), p=0.037] had higher odds of death. Patients who were admitted in the second week had lower odds of death and those admitted in the third week had higher odds of death.

**Interpretation:** This is the largest cohort of patients admitted with COVID-19 from India reported to date and has shown that vaccination status and early admission during the inflammatory phase can change the course of illness of these patients. Strategies should be made to improve vaccination rates and early admission of patients with moderate and severe COVID-19 to improve outcomes.

**Research in context:** *Evidence before this study:* The COVID-19 pandemic has been ravaging the world since December 2019 and the cases in various regions are being reported in waves. We found that the case fatality rates ranging from 1.4% to 28.3% have been reported in the first wave in India. Older age and the presence of comorbidities are known predictors of mortality. There are no reports regarding the effectiveness of vaccination, correlation of mortality with the timing of admission to the health care facility and inflammatory markers in the ‘second wave’ of the COVID-19 pandemic in India.

*Added-value of this study:* This study reports the real-world situation where patients get admitted at varying time points of their illness due to the mismatch between the availability of hospital beds and the rising number of COVID-19 patients during the pandemic. It reports the odds of developing severe hypoxia necessitating oxygen therapy and death thus helping identify priority groups for admission.

*Implications of all the available evidence:* This study found increased odds of requiring oxygen support or death in patients older than 45 years of age, with comorbidities, and those who had hyper-inflammation with raised C-reactive peptide, d-dimer or leukocytosis. Patients who were admitted in the second week of illness had lower odds of death as compared to those admitted in the third week implying that treatment with corticosteroids in the second week of the illness during the ‘inflammatory phase’ could lead to reduced mortality. These findings would help triage patients and provide guidance for developing admission policy during times where hospital beds are scarce. Vaccination was found to reduce the odds of deterioration or death and should be fast-tracked to prevent further ‘waves’ of the pandemic.

## Introduction

The SARS-CoV-2 infection has been declared a pandemic by the world health organization. To date, more than 200 million people have been infected by the virus and more than 4.26 million have died.^1^

The disease manifestation ranges from being completely asymptomatic and being detected only on screening due to a history of contact or may present with rapidly progressive hypoxic respiratory failure due to pneumonia and acute respiratory distress syndrome (ARDS).

The clinical features, demographic profile, severity at baseline and the case fatality rates differ between various geographic regions. We present these features and the predictors of outcomes in the Indian scenario during the ‘second wave’ of the COVID-19 pandemic in patients who were admitted to AIIMS, Jhajjar COVID-19 treatment facility. In the period from April to June 2021, this part of India was significantly affected by COVID-19 with many patients with nearly 30 percent positivity rates of tests conducted. The objectives of this study were to characterize the demographic, clinical, laboratory and imaging features, treatment trends and hospital outcomes in patients SARS-CoV-2 infection and to study the factors determining the outcomes in patients with SARS-CoV-2 infection.

## Methods

This was a retrospective cohort study conducted at a tertiary care teaching institute at the National Cancer Institute, All India Institute of Medical Sciences (AIIMS), Jhajjar, India. The study protocol was approved by the Institutional review board. The protocol was designed keeping in mind the STROBE checklist for observational studies. The National Cancer Institute, AIIMS, Jhajjar is a dedicated oncology center that was converted into a designated COVID-19 treatment facility. This referral institute catered to a wide area of the northern part of India and COVID-19 patients were referred here from Delhi as well as from the nearby states of Haryana, Punjab, Rajasthan Uttar Pradesh and Bihar.

From the hospital electronic database, we included all consecutive patients admitted from the COVID-19 screening area of the hospital. We retrospectively abstracted the clinical data using a structured data capture form from the case files, screening forms and treatment sheets. Laboratory parameters and results of other biochemical & microbiological reports were obtained from the hospital’s electronic patient information portal. The demographic parameters like age, gender, comorbidity status, vaccination status, vital parameters including oxygen saturation at presentation, treatment administered, course during the hospital stay, lab parameters at baseline and during the hospital stay and the outcomes in terms of discharge or death at end of hospitalization, were collected. June 21 was considered to be the cutoff date to calculate case fatality rates.

### Case definitions

#### SARS-CoV-2 infection

Patients with SARS-CoV-2 RNA detected on throat swab by RT-PCR or NAAT or SARS-CoV-2 antigen detected on Rapid antigen test.

#### Hypoxia

Any patient with oxygen saturation less than 94% on room air or needing oxygen >21% to maintain saturation on a ventilator was considered to be hypoxic.

#### COVID-Pneumonia

Patients with SARS-CoV-2 infection as described above with breathlessness and chest infiltrates on chest Xray or CT scan of the chest

#### COVID-19 ARDS

Patients with SARS-CoV-2 pneumonia as described above with symptoms and hypoxia developing in 7 or fewer days from onset along with ARDS as per Berlin Definition 2012.^2^

### COVID-19 severity^3^

#### Asymptomatic SARS-CoV-2 infection

Patients without symptoms of COVID-19 and positive for SARS-CoV-2 as described above.

#### Mild COVID-19

Patients with baseline oxygen saturation ≥94% without breathlessness but with other symptoms suggestive of COVID-19 such as fever, sore throat, myalgia, fatigue etc.

#### Moderate COVID-19

Patients with breathlessness and other symptoms suggestive of COVID-19 as described above, and with oxygen saturation ≥94%

#### Severe COVID-19

Patients with COVID-19 symptoms as described above with an oxygen saturation <94% or PaO2/FiO2<300 or respiratory rate >30/min.

### Renal dysfunction

Biochemistry report with a creatinine>1 mg/dL during the hospital stay or reduced urine output less than 0.5 mL per kg per hour or <400 mL per day or requiring hemodialysis for metabolic acidosis, hyperkalemia or encephalopathy due to renal dysfunction, as described above.

### Hospital-acquired infection

Biological samples from tracheal aspirates, urine, or blood cultures showing pathogens known to be associated with nosocomial infections.

#### Deteriorated during hospital stay

Patients who were not hypoxic at presentation, but went on to develop hypoxia; those who were on a face mask or non-rebreather mask receiving oxygen who went on to need high flow oxygen devices, non-invasive or invasive mechanical ventilation or those who needed renal replacement therapy for acute kidney injury which developed during the course of COVID-19.

#### Critical Illness

Development of respiratory failure necessitating mechanical ventilation, hypotension necessitating vasopressor support or renal dysfunction necessitating renal replacement therapy

#### Death

Patients who died due to any cause during the hospital stay

#### Death due to COVID-19

Death in which COVID-19 is the proximate or underlying cause of death according to the International guidelines for certification and classification (coding) of COVID-19as cause of death^4^

#### COVID-19 associated death

Cases where the associated COVID-19 infection could have aggravated the consequences of the primary illness or accident leading to death according to the International guidelines for certification and classification (coding) of COVID-19.^4^

#### Discharge

Persons with SARS-CoV-2 infection who were discharged alive from the hospital. This includes those who were discharged home, those who left against medical advice and those who were transferred to another medical facility as described below.

##### Discharged home

Patients with SARS-CoV-2 infection who were discharged alive from the hospital after recovery and the destination from the hospital was home.

##### Left against medical advice

Patients with SARS-CoV-2 infection who were discharged alive from the hospital before reaching the discharged home criteria as described above. Such patients may continue their treatment at another hospital or choose to go home.

##### Transfer

Patients with SARS-CoV-2 infection who were discharged alive from the hospital to another health care facility to allow continued medical care for their primary illness or COVID-19 related complications.

The data collected for the purpose of the study were de-identified and analyzed. The patients included in this analysis will also be used in other reports to study subgroups and to answer other research questions.

## Statistical analysis

The data were summarized using means and standard deviations for normal data and medians and interquartile ranges (p25-p75) for non-parametric data and means were compared using the ‘t test’ and medians using the Wilcoxon rank-sum test. The categorical data were summarized as proportions and compared using the Chi2 test or Fisher’s exact as appropriate. All statistical tests were performed with the use of a two-sided type I error rate of 5%. Missing data were not imputed and the summary parameters were calculated with the available data and the denominators (n) for each parameter was mentioned.

Univariate analysis was done to compare the various parameters between those who were discharged and those who died. Multivariate logistic regression analysis was done with models developed by including those that were found to be significant on univariate analysis as well as parameters of clinical relevance. We also included those parameters which we thought would influence the outcomes based on available scientific literature. Sensitivity analysis was done by dropping such parameters and by comparing the various models obtained by dropping them. Kaplan Meier survival probabilities were estimated by baseline severity status and were compared. All analysis was performed using STATA-Version 13.0 software.

## Results

A total of 2080 patients were admitted to our COVID-19 facility during the period from April – June 2021. Amongst these, 17 were admitted as caregivers of the patients and 46 were still admitted at the facility as of the cut-off date of the study. Figure 1 shows the sample recruitment for the analysis of this study.

**Figure 1:**
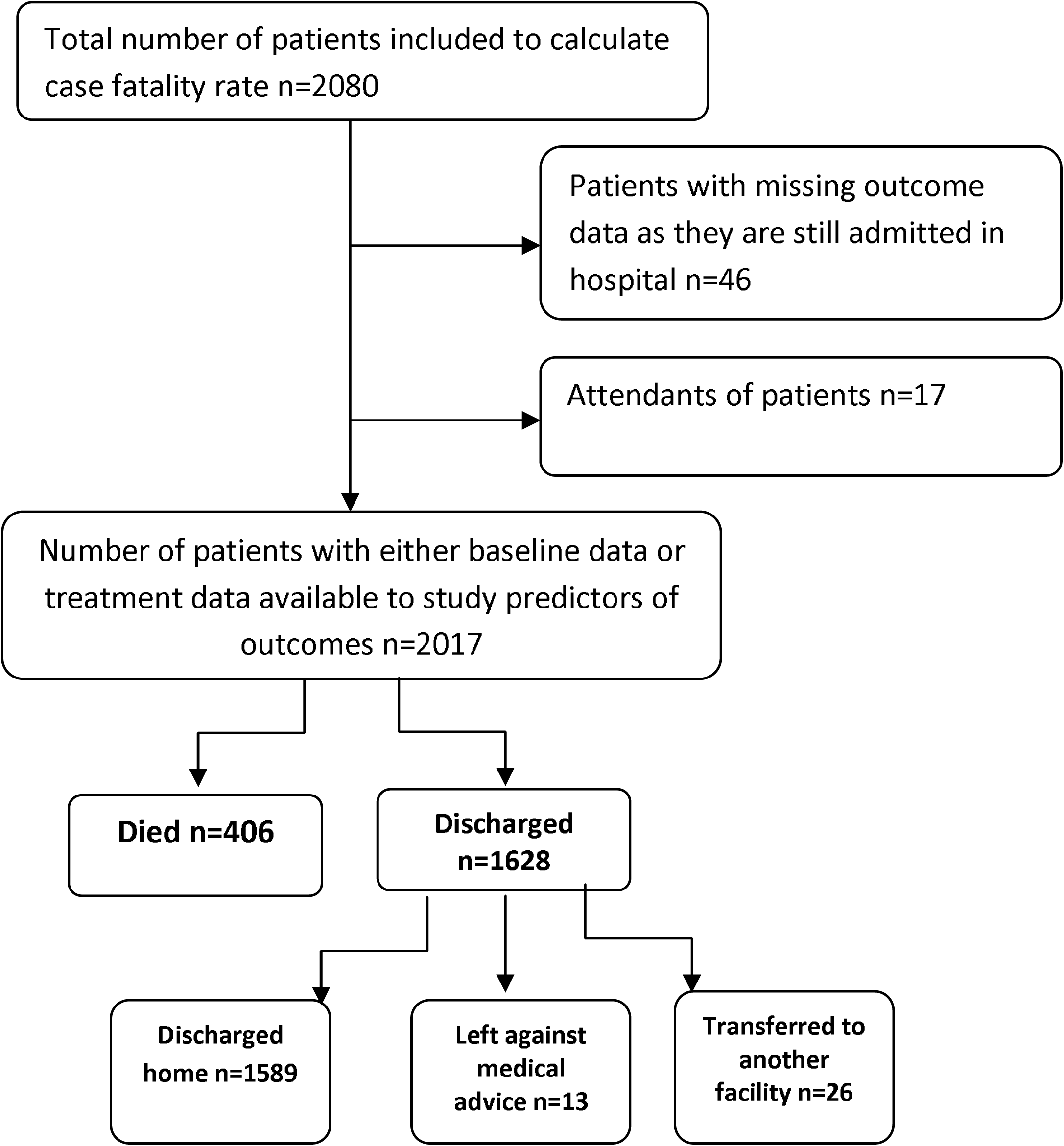
Patient recruitment and outcomes

### Case fatality rate and cause of death

Out of the 2080 admitted patients, there were 406 deaths, which amounted to a case fatality rate of 19.5%. In our cohort, 89 (21.9%) patients died less than 48 hours after admission. The majority of patients 342(84.2%) died due to refractory hypoxia. We had 28 patients (6.9%) who died due to myocarditis or sudden cardiac events such as pulmonary thromboembolism or myocardial infarction. Three patients were dead when they arrived at the emergency department. A minority of patients had acute kidney injury (5/406) and chronic kidney disease and uremia (2/406) as the cause of death. The other proximate causes of death were fall with head injury in one patient, SLE and related complications in two patients, febrile neutropenia and other complications of malignancy or chemotherapy (COVID-19 associated deaths) in eight patients, fungal pneumonia in 2 patients and mucor with fungal sepsis in four patients. The time to death and fraction of patients who were discharged at each time point has been categorized based on the severity of COVID-19 at presentation and the unadjusted odds are depicted in Figure 2.

**Figure-2:**
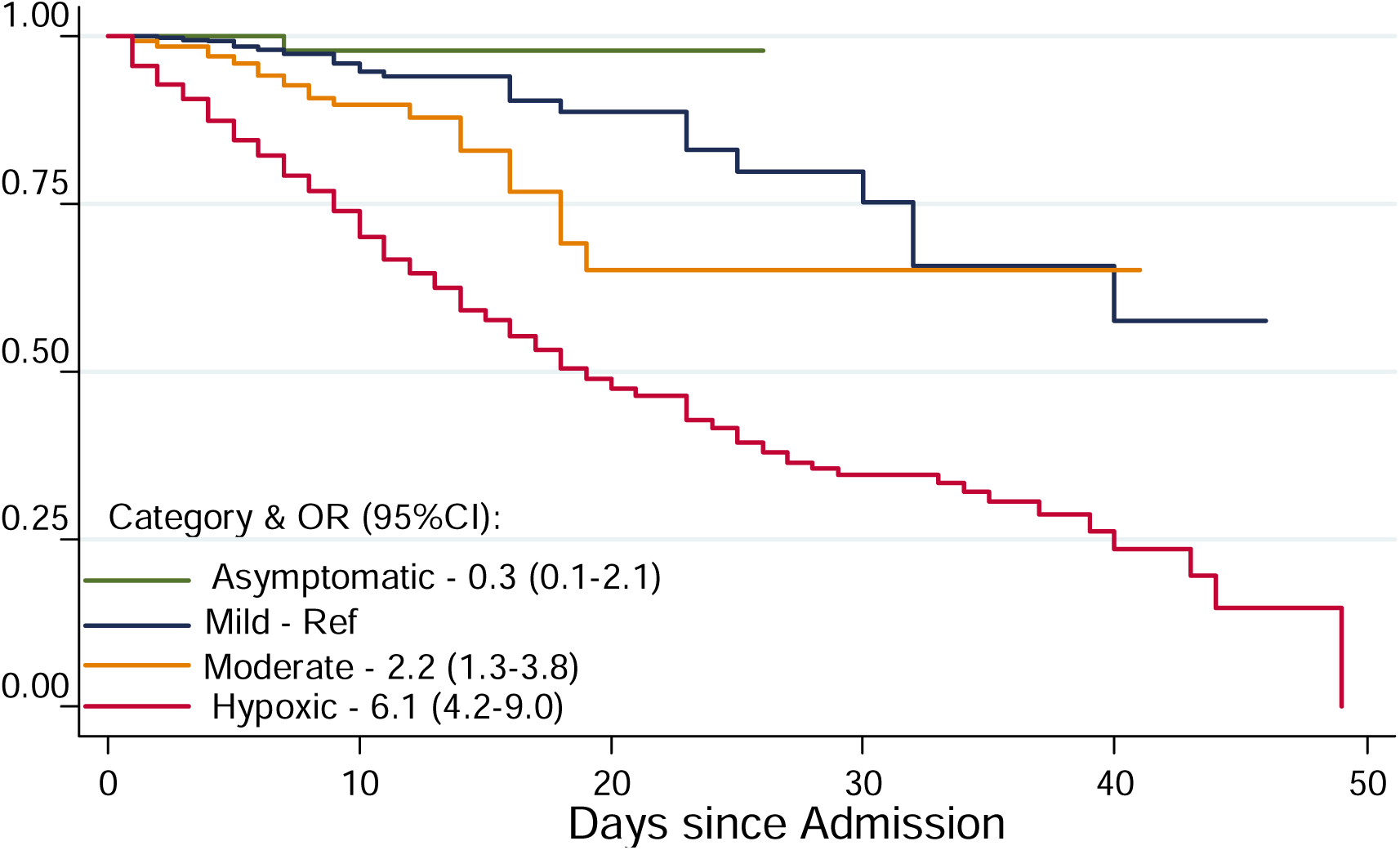
Kaplan-Meier survival estimates by COVID-19 baseline severity. Asymptomatic-Patients without symptoms of COVID-19 Mild-Patients with symptoms of COVID-19, but no breathlessness, SpO2 at baseline ≥94% Moderate-Patients with COVID-19 pneumonia and breathlessness, SpO2 at baseline ≥94% Severe-Patients with COVID-19 and severe hypoxia, SpO2 at baseline <94%

### Clinical features and baseline characteristics

Table 1 shows the sample characteristics of the 2017 patients who form the analysis cohort (Figure-1). Since this was a retrospective cohort study, we had missing data and it has been mentioned as to what proportion of the data points were missing for each of the parameters. 1763 (87.3%) patients were admitted due to COVID-19 symptoms and 99 (4.9%) patients were admitted for primarily non-COVID-19 indications such as malignancy, pancreatitis etc. and were found to be positive for SARS-CoV-2 during routine testing during the hospital stay. In our cohort, males 1355(65%) outnumbered females 725 (35%). We had 1572 patients who were discharged from the hospital cured or improved, 13 patients who left against medical advice and 26 patients who were transferred to other medical facilities for continuing care.

**Table 1:**
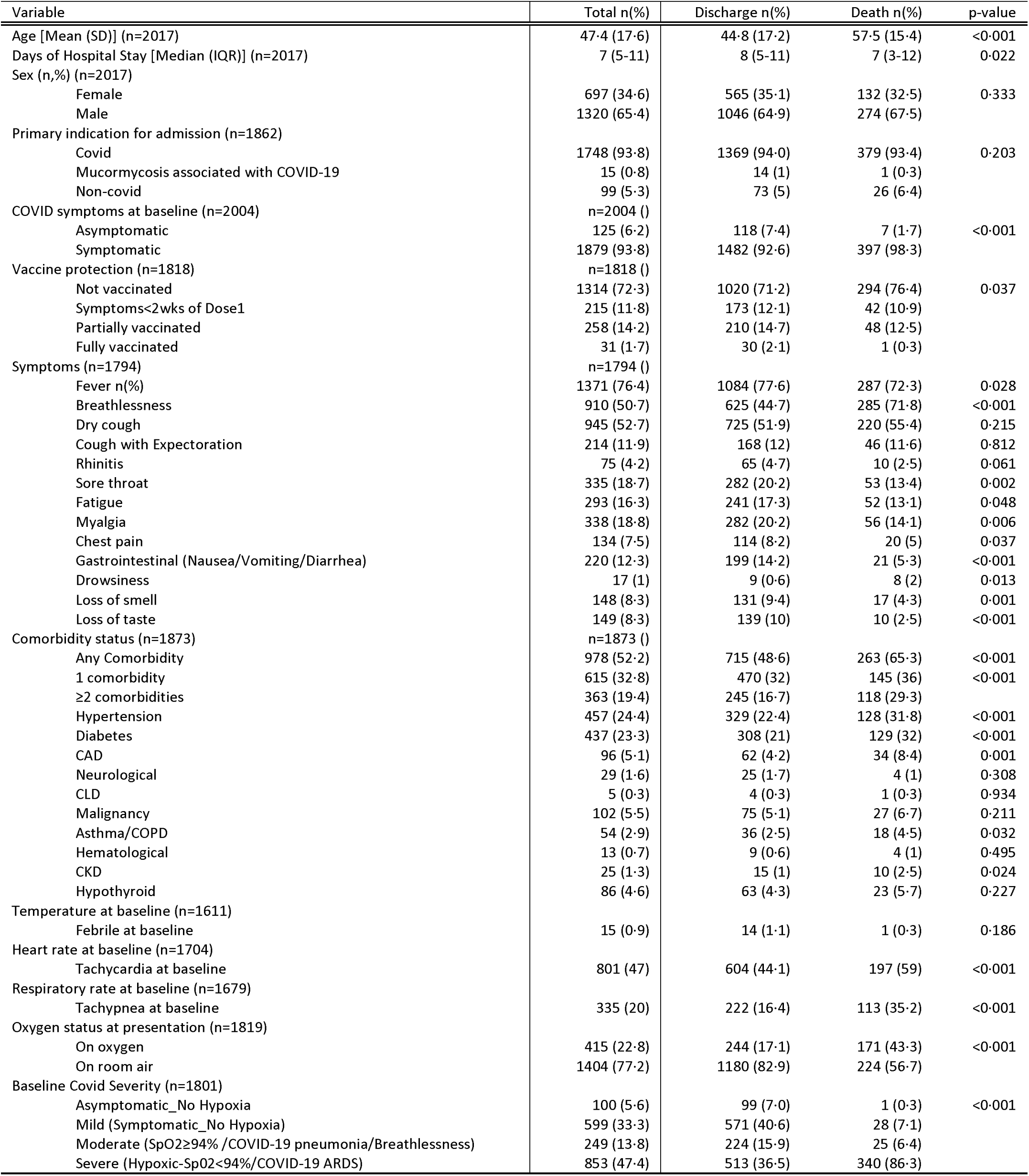
Demographic and clinical profile of patients at baseline

| Variable | Total n(%) | Discharge n(%) | Death n(%) | p-value |
| --- | --- | --- | --- | --- |
| Age [Mean (SD)] (n=2017) | 47.4 (17.6) | 44.8 (17.2) | 57.5 (15.4) | <0.001 |
| Days of Hospital Stay [Median (IQR)] (n=2017) | 7 (5-11) | 8 (5-11) | 7 (3-12) | 0.022 |
| Sex (n,%) (n=2017) |  |  |  |  |
| Female | 697 (34.6) | 565 (35.1) | 132 (32.5) | 0.333 |
| Male | 1320 (65.4) | 1046 (64.9) | 274 (67.5) |  |
| Primary indication for admission (n=1862) |  |  |  |  |
| Covid | 1748 (93.8) | 1369 (94.0) | 379 (93.4) | 0.203 |
| Mucormycosis associated with COVID-19 | 15 (0.8) | 14 (1) | 1 (0.3) |  |
| Non-covid | 99 (5.3) | 73 (5) | 26 (6.4) |  |
| COVID symptoms at baseline (n=2004) | n=2004 ( ) |  |  |  |
| Asymptomatic | 125 (6.2) | 118 (7.4) | 7 (1.7) | <0.001 |
| Symptomatic | 1879 (93.8) | 1482 (92.6) | 397 (98.3) |  |
| Vaccine protection (n=1818) | n=1818 ( ) |  |  |  |
| Not vaccinated | 1314 (72.3) | 1020 (71.2) | 294 (76.4) | 0.037 |
| Symptoms<2wks of Dose1 | 215 (11.8) | 173 (12.1) | 42 (10.9) |  |
| Partially vaccinated | 258 (14.2) | 210 (14.7) | 48 (12.5) |  |
| Fully vaccinated | 31 (1.7) | 30 (2.1) | 1 (0.3) |  |
| Symptoms (n=1794) | n=1794 ( ) |  |  |  |
| Fever n(%) | 1371 (76.4) | 1084 (77.6) | 287 (72.3) | 0.028 |
| Breathlessness | 910 (50.7) | 625 (44.7) | 285 (71.8) | <0.001 |
| Dry cough | 945 (52.7) | 725 (51.9) | 220 (55.4) | 0.215 |
| Cough with Expectoration | 214 (11.9) | 168 (12) | 46 (11.6) | 0.812 |
| Rhinitis | 75 (4.2) | 65 (4.7) | 10 (2.5) | 0.061 |
| Sore throat | 335 (18.7) | 282 (20.2) | 53 (13.4) | 0.002 |
| Fatigue | 293 (16.3) | 241 (17.3) | 52 (13.1) | 0.048 |
| Myalgia | 338 (18.8) | 282 (20.2) | 56 (14.1) | 0.006 |
| Chest pain | 134 (7.5) | 114 (8.2) | 20 (5) | 0.037 |
| Gastrointestinal (Nausea/Vomiting/Diarrhea) | 220 (12.3) | 199 (14.2) | 21 (5.3) | <0.001 |
| Drowsiness | 17 (1) | 9 (0.6) | 8 (2) | 0.013 |
| Loss of smell | 148 (8.3) | 131 (9.4) | 17 (4.3) | 0.001 |
| Loss of taste | 149 (8.3) | 139 (10) | 10 (2.5) | <0.001 |
| Comorbidity status (n=1873) | n=1873 ( ) |  |  |  |
| Any Comorbidity | 978 (52.2) | 715 (48.6) | 263 (65.3) | <0.001 |
| 1 comorbidity | 615 (32.8) | 470 (32) | 145 (36) | <0.001 |
| ≥2 comorbidities | 363 (19.4) | 245 (16.7) | 118 (29.3) |  |
| Hypertension | 457 (24.4) | 329 (22.4) | 128 (31.8) | <0.001 |
| Diabetes | 437 (23.3) | 308 (21) | 129 (32) | <0.001 |
| CAD | 96 (5.1) | 62 (4.2) | 34 (8.4) | 0.001 |
| Neurological | 29 (1.6) | 25 (1.7) | 4 (1) | 0.308 |
| CLD | 5 (0.3) | 4 (0.3) | 1 (0.3) | 0.934 |
| Malignancy | 102 (5.5) | 75 (5.1) | 27 (6.7) | 0.211 |
| Asthma/COPD | 54 (2.9) | 36 (2.5) | 18 (4.5) | 0.032 |
| Hematological | 13 (0.7) | 9 (0.6) | 4 (1) | 0.495 |
| CKD | 25 (1.3) | 15 (1) | 10 (2.5) | 0.024 |
| Hypothyroid | 86 (4.6) | 63 (4.3) | 23 (5.7) | 0.227 |
| Temperature at baseline (n=1611) |  |  |  |  |
| Febrile at baseline | 15 (0.9) | 14 (1.1) | 1 (0.3) | 0.186 |
| Heart rate at baseline (n=1704) |  |  |  |  |
| Tachycardia at baseline | 801 (47) | 604 (44.1) | 197 (59) | <0.001 |
| Respiratory rate at baseline (n=1679) |  |  |  |  |
| Tachypnea at baseline | 335 (20) | 222 (16.4) | 113 (35.2) | <0.001 |
| Oxygen status at presentation (n=1819) |  |  |  |  |
| On oxygen | 415 (22.8) | 244 (17.1) | 171 (43.3) | <0.001 |
| On room air | 1404 (77.2) | 1180 (82.9) | 224 (56.7) |  |
| Baseline Covid Severity (n=1801) |  |  |  |  |
| Asymptomatic_No Hypoxia | 100 (5.6) | 99 (7.0) | 1 (0.3) | <0.001 |
| Mild (Symptomatic_No Hypoxia) | 599 (33.3) | 571 (40.6) | 28 (7.1) |  |
| Moderate (SpO2≥94% /COVID-19 pneumonia/Breathlessness) | 249 (13.8) | 224 (15.9) | 25 (6.4) |  |
| Severe (Hypoxic-SpO2<94%/COVID-19 ARDS) | 853 (47.4) | 513 (36.5) | 340 (86.3) |  |

We found that around half of the patients (47%) had severe disease at presentation (oxygen saturation of <94% at baseline). Tachycardia, tachypnea and hypoxia at presentation were associated with increased odds of mortality on univariate analysis. Patients who had breathlessness and those who were drowsy or in altered sensorium had a higher odds of death. Sore throat, fatigue, myalgia, chest pain, gastrointestinal symptoms, loss of smell and taste were reported in higher proportions by patients who survived. This is likely due to the higher proportion of patients with severe disease, who probably could not narrate the history due to their respiratory distress. Breathlessness and altered sensorium on the other hand can be assessed by the examiner and could have been more accurately captured thus attaining significant differences between those who survived and those who died. Of note, 13.8% of patients presented with COVID-19 pneumonia and 47.4% of patients presented with COVID-19 related ARDS.

### Laboratory parameters

The biochemical parameters of the cohort at baseline are detailed in Table 2. We found that patients with hyper inflammation as evidenced by leucocytosis, elevated C-reactive peptide and d-dimer were associated with higher odds of death.

**Table 2:** Laboratory parameters at baseline

| Parameter | Discharges |  | Deaths |  | p value* |
| --- | --- | --- | --- | --- | --- |
|  | Available observations | Mean(SD) or Median (p25-p75) | Available observations | Mean(SD) or Median (p25-p75) |  |
| Hemoglobin (mg/dL) | 1322 | 12.9 (2.2) | 334 | 12.7 (2.2) | 0.051 |
| Leucocyte count (cells/mm<sup>3</sup>) | 1320 | 6.8 (4.8-9.9) | 332 | 11.6 (8.1-15.9) | <0.001 |
| Neutrophil (cells/mm<sup>3</sup>) | 1287 | 4.8 (2.8-7.8) | 320 | 10.0 (6.6-13.9) | <0.001 |
| Lymphocytes (cells/mm<sup>3</sup>) | 1288 | 1.0 (0.7-1.5) | 320 | 0.6 (0.4-0.8) | <0.001 |
| LDH (U/L) | 1159 | 328 (241-460) | 271 | 654 (501-890) | <0.001 |
| T- Bilirubin (mg/dL) | 1341 | 0.5 (0.4-0.7) | 339 | 0.6 (0.4-0.9) | <0.001 |
| ALT (U/L) | 1329 | 45 (27-79) | 338 | 53 (31-91) | 0.003 |
| AST (U/L) | 1329 | 42 (30-68) | 338 | 61 (39-98) | <0.001 |
| Globulin (g/dL) | 1315 | 2.6 (0.4) | 336 | 2.7 (0.5) | 0.002 |
| A/G ratio | 1314 | 1.5 (1.4-1.7) | 334 | 1.3 (1.1-1.5) | <0.001 |
| Albumin (g/dL) | 1329 | 3.9 (0.5) | 337 | 3.5 (0.5) | <0.001 |
| Urea (mg/dL) | 1351 | 30 (21.4-43.0) | 338 | 59.9 (42.8-87.7) | <0.001 |
| Creatinine (mg/dL) | 1351 | 0.7 (0.6-0.9) | 338 | 0.9 (0.7-1.3) | <0.001 |
| Calcium (mg/dL) | 1323 | 8.6 (3.6) | 338 | 8.1 (0.8) | <0.001 |
| Phosphate (mg/dL) | 1323 | 3.0 (2.5-3.5) | 338 | 3.1 (2.5-4.0) | 0.005 |
| Sodium (mEq/L) | 1341 | 137.2 (8.3) | 339 | 139.2 (9.7) | <0.001 |
| Potassium (mEq/L) | 1341 | 4.5 (4.1-4.9) | 339 | 4.7 (4.2-5.2) | <0.001 |
| *Inflammatory markers* |  |  |  |  |  |
| • Ferritin (ng/mL) | 473 | 365.4 (135.2-878) | 116 | 1096.5 (712.5-1650) | <0.001 |
| • D-Dimer (ng/mL) | 1154 | 201.5 (121-347) | 271 | 622 (337-2550) | <0.001 |
| • CRP (mg/dL) | 1286 | 2.6 (0.8-7.4) | 301 | 9.0 (5.1-16.0) | <0.001 |
| • IL-6 (pg/mL) | 581 | 9.1 (3.4-22.6) | 195 | 40.5 (14.4-104.2) | <0.001 |
| **Categorical** | **n** | **n( %)** | **n** | **n( %)** | **chi2-pvalue** |
| *Hemoglobin<12g/dl* | 1322 | 378 (28.6) | 334 | 111 (33.2) | 0.097 |
| *Thrombocytopenia(<1.5Lac)* | 1329 | 269 (20.4) | 334 | 74 (22.2) | 0.439 |
| Leucopenia | 1320 | 199 (15.1) | 332 | 15 (4.5) | <0.001 |
| LDH>246U/L | 1159 | 853(73.6) | 271 | 269 (99.3) | <0.001 |
| Total Bilirubin>1.2mg/dl | 1341 | 60 (4.5) | 339 | 29 (8.6) | 0.003 |
| ALT >49U/L | 1329 | 605 (45.5) | 338 | 179 (53.0) | 0.014 |
| AST>=34U/L | 1329 | 889 (66.9) | 338 | 275 (81.4) | <0.001 |
| Urea >=50mg/dl | 1351 | 242 (17.9) | 338 | 213 (63.0) | <0.001 |
| Creatinine>=1.0mg/dl | 1351 | 53 (3.9) | 338 | 88 (26.0) | <0.001 |
| Hyperferritinemia (>322 ng/mL) | 473 | 250 (52.9) | 116 | 105 (90.5) | <0.001 |
| d-dimer >500 ng/ml | 1154 | 188 (16.3) | 271 | 155 (57.2) | <0.001 |
| IL-6 >4.4 pg/ml | 581 | 410 (70.6) | 195 | 188 (95.4) | <0.001 |
| CRP >0.5 mg/dl | 1286 | 1031 (80.2%) | 301 | 298 (99.0%) | <0.001 |

Notes: p-values for means are calculated by t-test and for medians by Wilcoxon rank-sum test.

### Treatment trends

The patients were treated by the team of intensive care physicians according to the clinical condition based on the national guidelines, institute protocols and latest scientific literature subject to availability of the interventions. In summary, almost all patients who were hypoxic and were on oxygen were treated with corticosteroids. All hypoxic patients without significant thrombocytopenia or other clear contraindications were treated with prophylactic doses of anticoagulation. The proportion of patients who were discharged and those who expired receiving various medications is detailed in Table 3. As expected, patients with severe disease were more likely to be treated with drugs such as pulse methylprednisolone, tocilizumab and remdesivir. Out of 256 patients who were treated with non-invasive ventilation, 221(86.3%) succumbed while only 13.7% of patients survived. Similarly, 250 patients received invasive mechanical ventilation, thereby representing 13.7% of the total cohort. Only 2.4% of those who received mechanical ventilation could be discharged and the rest succumbed to the illness.

**Table 3:** Treatment offered and in-hospital complications

| Parameter (n) | Total<br>n (% of observations<br>who received the<br>intervention) | Discharged<br>n (%)* | Died<br>n (%)* | chi2/Exact<br>p value |
| --- | --- | --- | --- | --- |
| Oxygen n=1831 | 1025 (56.0) | 626 (61.1) | 399 (38.9) | <0.001 |
| HFNC n=1808 | 138 (7.6) | 42 (30.4) | 96 (69.6%) | <0.001 |
| NIV n=1823 | 256 (14.0) | 35 (13.7) | 221 (86.3%) | <0.001 |
| Invasive MV n=1830 | 250 (13.7) | 6 (2.4) | 244 (97.6) | <0.001 |
| ICU admission n=2017 | 239 (11.9) | 99 (41.4) | 140 (58.6) | <0.001 |
| Corticosteroid use (n=1813) | 1094 (60.3) | 722 (66.0) | 372 (34.0) | <0.001 |
| Pulse methylprednisolone n=1752 | 165 (9.4) | 52 (31.5) | 113 (68.5) | <0.001 |
| Inhaled steroids n=1761 | 357 (20.3) | 247 (69.2) | 110 (30.8) | <0.001 |
| Anticoagulation therapy n=1796 | 988 (55.0) | 658 (66.6) | 330 (33.4) | <0.001 |
| Ivermectin n=1758 | 283 (16.1) | 235 (83.0) | 48 (17.0) | 0.115 |
| Doxycycline n=1757 | 324 (18.4) | 246 (75.9) | 78 (24.1) | 0.072 |
| Minocycline n= 1757 | 38 (2.2) | 29 (76.3) | 9 (23.7) | 0.615 |
| Azithromycin n=1754 | 247 (14.1) | 195 (79.0) | 52 (21.1) | 0.750 |
| Ceftriaxone n=1757 | 286 (16.3) | 172 (60.1) | 114 (39.9) | <0.001 |
| Levofloxacin n=1760 | 234 (13.3) | 98 (41.9) | 136 (58.1) | <0.001 |
| Tocilizumab n=1755 | 37 (2.1) | 8 (21.6) | 29 (78.4) | <0.001 |
| Remdesivir n=1755 | 403 (23.0) | 257 (63.8) | 146 (36.2) | <0.001 |
| Zinc n=1687 | 486 (28.8) | 440 (90.5) | 46 (9.5) | <0.001 |
| Hyperglycemia n=1660 | 402 (24.2) | 258 (64.2) | 144 (35.8) | <0.001 |
| Renal dysfunction n=1831 | 340 (18.6) | 152 (44.7) | 188 (55.3) | <0.001 |
| Hypotension n=1800 | 95 (5.3) | 3 (3.2) | 92 (96.8) | <0.001 |
| Hemodialysis n=2017 | 20 (1.0) | 6 (30.0) | 14 (70.0) | <0.001 |
| Dialysis for AKI | 5 (0.25) | 1 (20.0) | 4 (80.0) | <0.001 |

\*n denotes the number of patients who received the intervention and % denotes the proportion of patients who were discharged or died after receiving the intervention

### Predictors of outcome

We analyzed the predictors of (i) developing severe illness needing oxygen therapy, (ii) deterioration during the hospital stay as defined above using the clinical and laboratory parameters at baseline are depicted in Tables 4 and 5 respectively. Predictors for the development of critical illness (as defined above) are depicted in supplementary table-1. The independent predictors of death were derived using logistic regression analysis and 4 models were created using the clinically relevant parameters as covariates. Adjusting for baseline clinical characteristics (Table-6, Model-1), age>=45years, having comorbid conditions, Severe illness at presentation and hospitalization in the 3^rd^ week of illness were independently associated with increased odds of death. Vaccination and hospitalization in the 2^nd^ week of illness reduced the odds of death by 30% (OR 0.7 95% CI 0.5-0.9 p-value 0.008) and 36% (OR 0.64 95% CI 0.48-0.86 p-value 0.003) respectively. The factors predicting deterioration in patients who were not hypoxic at baseline have been described in Supplementary table-2.

**Table 4:** Factors predicting the development of hypoxia requiring Oxygen Support

Oxygen Support during the hospital stay
|  |  | Model-1 | Model-2 |
| --- | --- | --- | --- |
|  |  | OR(95%CI), p-value | OR(95%CI), p-value |
| Age (Ref: 18-45yrs) |  |  |  |
|  | <18 years | **0.5 (0.2-0.9), 0.035** | **0.2 (0.1-0.8), 0.022** |
|  | 45-60 years | **2.8 (2.1-3.8), <0.001** | **1.9(1.3-2.8), 0.001** |
|  | >60yrs | **2.8 (2-4), <0.001** | **1.6 (1.0-2.6), 0.044** |
| Male (Ref: Female) |  | 1.2 (1.0-1.6), 0.094 | 1.0 (0.7-1.5), 0.846 |
| Vaccinated |  | **0.6 (0.5-0.8), <0.001** | **0.7 (0.5-0.9), 0.047** |
| Symptom onset to Hospitalization (Ref: 1week) |  |  |  |
|  | 2weeks | 1.2 (0.9-1.5), 0.248 | 1.0 (0.7-1.4), 0.970 |
|  | 3 or more weeks | 1.9 (0.47-8.1), 0.362 | 2.3 (0.3-20), 0.463 |
|  | Asymptomatic | **0.3 (0.2-0.6), <0.001** | **0.1 (0.04-0.3), <0.001** |
| Symptoms reported (Ref: No) |  |  |  |
|  | Breathlessness | **6.4 (5-8.2), <0.001** | **3.7 (2.7-5.2), <0.001** |
|  | Dry cough | **1.3 (1-1.7), 0.021** |  |
|  | Rhinitis | **0.5 (0.3-0.8), 0.012** |  |
|  | Sore throat | **0.7 (0.5-0.9), 0.021** |  |
|  | Fatigue | **0.7 (0.5-0.9), 0.037** |  |
| Comorbidities (Ref: No) |  |  |  |
|  | 1 | **1.4 (1-1.8), 0.032** | 1.3 (0.9-1.9), 0.171 |
|  | 2 or more | **1.7 (1.2-2.5), 0.003** | 1.6 (1.0-2.6), 0.055 |
| Lab Parameters (Ref: No) |  |  |  |
| Leucocyte Count (Ref: Normal) |  |  |  |
|  | Leukopenia |  | **0.4 (0.2-0.6), <0.001** |
|  | Leukocytosis |  | **2.8 (1.8-4.3), <0.001** |
| D-Dimer>500 (Ref:≤500) |  |  | **2.5 (1.6-3.8), <0.001** |
| CRP high (Ref: Normal) |  |  | **5.7 (3.4-9.7), <0.001** |
| Creatinine>1.0 mg/dl |  |  | **1.7 (1.1-2.8), 0.024** |

Notes: Model-1: Adjusting for baseline clinical parameters, Model-2: Model-1 plus baseline lab parameters. Age, gender and comorbidities are included in all models. Only symptoms and lab parameters with significant p-value are included in these final models.

**Table 5:** Factors predicting deterioration during the hospital stay

| Deterioration during the hospital stay |  |  |
| --- | --- | --- |
|  | Model-1 | Model-2 |
|  | OR(95%CI), p-value | OR(95%CI), p-value |
| Age (Ref: 18-45yrs) |  |  |
| <18 years | 0.6 (0.3-1.4), 0.217 | 0.5 (0.1-2.0), 0.336 |
| 45-60 years | **1.8 (1.3-2.4), <0.001** | 1.5 (1.0-2.2), 0.040 |
| >60years | **2.6 (1.9-3.6), <0.001** | 1.8 (1.2-2.7), 0.008 |
| Male (Ref: Female) | 1.2 (0.9-1.5), 0.168 | 0.9 (0.7-1.3), 0.660 |
| Primary Condition (Ref: Covid) |  |  |
| Non-Covid | **2.1 (1.2-3.7), 0.012** | 0.9 (0.4-2.2), 0.825 |
| mucor | 1.7 (0.1-20.4), 0.686 |  |
| Vaccinated | 0.8 (0.6-1.0), 0.067 | 0.8 (0.6-1.1), 0.140 |
| Time to Admission from Symptom Onset (Ref: 1wk) |  |  |
| 2weeks | **0.7 (0.5-0.8), 0.001** | **0.6 (0.4-0.8), 0.001** |
| 3 or more weeks | **3.1 (1.1-9.6), 0.046** | 2.0 (0.4-10.0), 0.412 |
| Asymptomatic | 1.0 (0.3-3.7), 0.948 | 2.3 (0.1-39.0), 0.572 |
| Comorbidities (Ref: No) |  |  |
| 1 | **1.3 (1.02-1.7), 0.037** | 1.3 (0.9-1.9), 0.107 |
| 2 or more | **1.6 (1.2-2.2), 0.004** | 1.5 (1.0-2.2), 0.078 |
| Baseline COVID-19 severity (Ref: Mild) |  |  |
| Asymptomatic – No hypoxia | **0.2 (0.1-0.8), 0.026** | 0.1 (0.0-2.2), 0.146 |
| Moderate | **2.4 (1.7-3.5), <0.001** | **1.6 (1.0-2.6), 0.032** |
| Severe | **3.0 (2.3-4.0), <0.001** | 1.1 (0.7-1.6), 0.697 |
| Symptoms reported (Ref: No) |  |  |
| Fever | **0.7 (0.5-0.9), 0.020** |  |
| Dry cough | **1.3 (1.1-1.7), 0.020** | **1.6 (1.2-2.2), 0.002** |
| Lab Parameters (Ref: No) |  |  |
| Thrombocytopenia |  | **1.8 (1.2-2.6), 0.004** |
| Leucocyte Count (Ref: Normal) |  |  |
| Leukopenia |  | **0.5 (0.3-0.8), 0.005** |
| Leukocytosis |  | **2.1 (1.5-2.9), <0.001** |
| D-Dimer>500 (Ref:≤500) |  | **2.5 (1.8-3.4), <0.001** |
| CRP high (Ref: Normal) |  | **6.0 (2.8-13.0), <0.001** |
| Creatinine>1.0 mg/dl |  | **2.2 (1.5-3.2), <0.001** |

Notes: Model-1: Adjusting for baseline clinical parameters, Model-2: Model-1 plus baseline lab parameters. Age, gender and comorbidities are included in all models. Only symptoms and lab parameters with significant p-value are included in these final models.

**Table 6:**
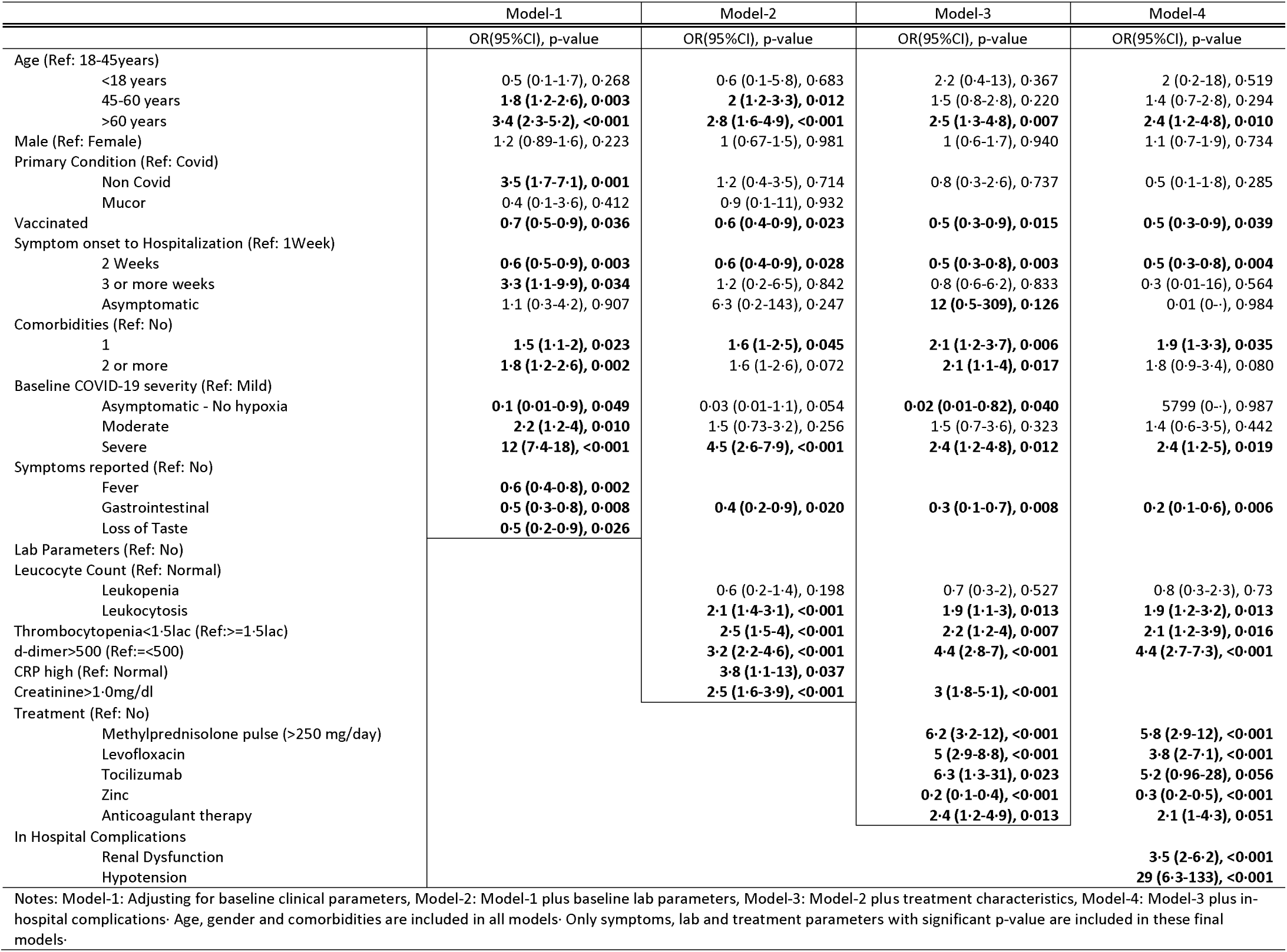
Factors predicting death

Leukocytosis, thrombocytopenia, elevated d-dimer, C-reactive peptide and creatinine at baseline independently predicted mortality after adjusting for other baseline and treatment characteristics. (Table-6, Model-3).

## Discussion

This study describes the demography and clinical profile of the patients admitted to our facility during the ‘second wave’ of the COVID-19 pandemic. Few points are worthy of elaboration. Though the vaccination program had been started around 3 months before the rapid ascent in the curve, less than 2% of the patients who were admitted had received both the doses of vaccine at least 2 weeks before the infection and 14.2% were partially vaccinated. The designated COVID-19 facility catered to predominantly patients who were getting admitted due to COVID-19 pneumonia. We had around 5% of patients who had other indications for admission such as malignancy. We found that getting admitted for other indications had a higher odds of death as compared to those who were admitted for COVID-19. Similarly, another interesting finding was that patients who got admitted in the third week [OR 3.3 95% CI 1.1-9.7 p=0.027] after symptom onset had a higher odds of death as compared to those getting admitted in the first week of onset of symptoms, while the odds of death was significantly lower [OR 0.6 95% CI 0.5-0.8 95% CI 0.001] when admitted during the second week of onset of symptoms compared to week-1. This probably could be because the initial first week was associated with fulminant viral pneumonia and those who present with severe disease at this early viraemic stage had a higher odds of death. Likewise, those who presented in the third week are likely to be those with inflammatory damage due to the ‘cytokine storm’. These patients who presented in the third week are likely those who had not received corticosteroid therapy in the second week of the illness when the inflammatory phase had set in, probably leading to an increase in mortality. However, this hypothesis needs further study.

Our findings were different from the study by Chauhan et al.^5^ that showed more proportion of fever, fatigue, myalgia, abdominal pain in those who died as compared to those who survived. However, the study by Bairwa et al.^6^ concurred with the clinical symptomatology of our study. The case fatality rate of our cohort was 19.5% which was similar to that in multiple other studies.^5–9^ Another cohort from our center during the ‘first wave’ of the pandemic reported a Case fatality rate of 1.4%.^10^ However a study from a tertiary care hospital in New Delhi reported a CFR of 28.3%.^11^The cohort of patients reported by Gupta et al.^12^ had a case fatality rate of 9.5% while the study by Wang et al.^13^ reported a CFR of 4.3%. These differences could be due to various factors such as different strains of the virus, vaccination rates, usage of antiviral/anti-inflammatory agents and monoclonal antibodies, due to study design factors such as the proportion of patients still admitted and undergoing treatment at the end of the study or due to other baseline differences. Similar to these studies, a higher proportion of those with comorbidities and those with severe illness succumbed to the illness. In a subset of patients from this study who were over 18 years of age, and were eligible for vaccination, it was found that those who had completed the course of vaccination had 86% reduced odds of developing hypoxia and had a case fatality rate of 5.6% as compared to 22.8% in the unvaccinated group.^14^

Patients treated with methylprednisolone pulse therapy, remdesivir and tocilizumab had a higher odds of death as reflected in Table 4. This apparent paradox may be due to confounding by indication. Understandably, patients with severe disease and those with critical illness are more likely to be treated with these agents.

In our cohort, we had 1067 patients who needed oxygen and only 63% of them survived. This implies a mortality rate of 37% in patients who needed some form of oxygen therapy or mechanical ventilation. This reflects the severe nature of COVID-19 pneumonia and ARDS in this cohort of patients. In a subset of patients of this cohort with hypoxia at presentation, it was found that the case fatality rate was 45.4% in those with silent hypoxia 40.03% in those with dyspneic hypoxia.^15^ More than 47% of patients of our cohort had ARDS due to COVID-19 pneumonia. Respiratory failure necessitating mechanical ventilation had a very poor prognosis with only 13.7% of those receiving non-invasive ventilation and 2.4% of those receiving invasive mechanical ventilation being discharged. It was disappointing that, almost 86% of patients who had to be ventilated died. This is probably due to the fulminant viral pneumonia in the initial week and hyperinflammatory lung disease during the second and third weeks of the illness. This is different from the historical mortality rates of ARDS.^16^ Similarly, all 12 patients who developed barotrauma-related pneumothorax or pneumomediastinum succumbed. The presence of renal dysfunction irrespective of the need for renal replacement therapy and hypotension were also independently associated with increased odds of mortality.

Our study includes a large cohort of patients which allows us to explore the various factors associated with severe illness, deterioration during hospitalization and death. However, it has a few limitations. Due to the retrospective nature of the data collection, few files had been misplaced or lost and nearly 10% of data points were missing. However, due to the large sample size of 2080 patients, we were able to find several independent risk factors for these outcomes. Similarly, due to the observational nature of the study, it is not possible to draw conclusions regarding causation. It is difficult to comment on the efficacy of for example high dose methylprednisolone pulses in preventing in-hospital deterioration because the drug had been administered at varying time points in the patients rather than a protocolized administration. In some patients, it had been administered once they needed oxygen administered through a high-flow nasal cannula while in others it was administered while on the verge of respiratory failure needing non-invasive ventilation. Similarly, the treatment was decided by the individual clinicians based on their discretion and thus subject to confounding by indication, thus precluding definite causation from being inferred. Other inflammatory markers such as raised ferritin and IL-6 were also associated with increased odds of death, however, due to the low number of patients in whom these parameters were available at baseline, it was not included in the logistic regression models.

## Conclusion

This is the largest cohort of COVID-19 patients that has been reported from the Southeast Asian region. Our study has also reported the relationship between the time of presentation and its association with mortality. We believe that early admission to the hospital, especially during the inflammatory phase, could make a difference by reducing mortality. Though during the first week or the viremic phase, we have no definite intervention to prevent mortality. Even remdesivir, which is an antiviral drug is effective in patients who are hypoxic but not critically ill and not in the early phase of the illness. Based on the available data, we could also conclude that vaccination has an impact on reducing the odds of death. However, once the patient develops ARDS related to COVID-19 necessitating respiratory support, the prognosis is dismal. Early administration of corticosteroids early in the inflammatory phase seems to be the only intervention that could have possibly changed the course of illness in our patients.

### Data sharing

data collected for the study, including individual participant data and a data dictionary defining each field in the set, will be made available to others upon reasonable request to be routed through our Institute ethics committee with an appropriate protocol

## Data Availability

Data sharing: data collected for the study, including individual participant data and a data dictionary defining each field in the set, will be made available to others upon reasonable request to be routed through our Institute ethics committee with an appropriate protocol

## Funding

No funding was received

---

**Supplementary Table 1:**
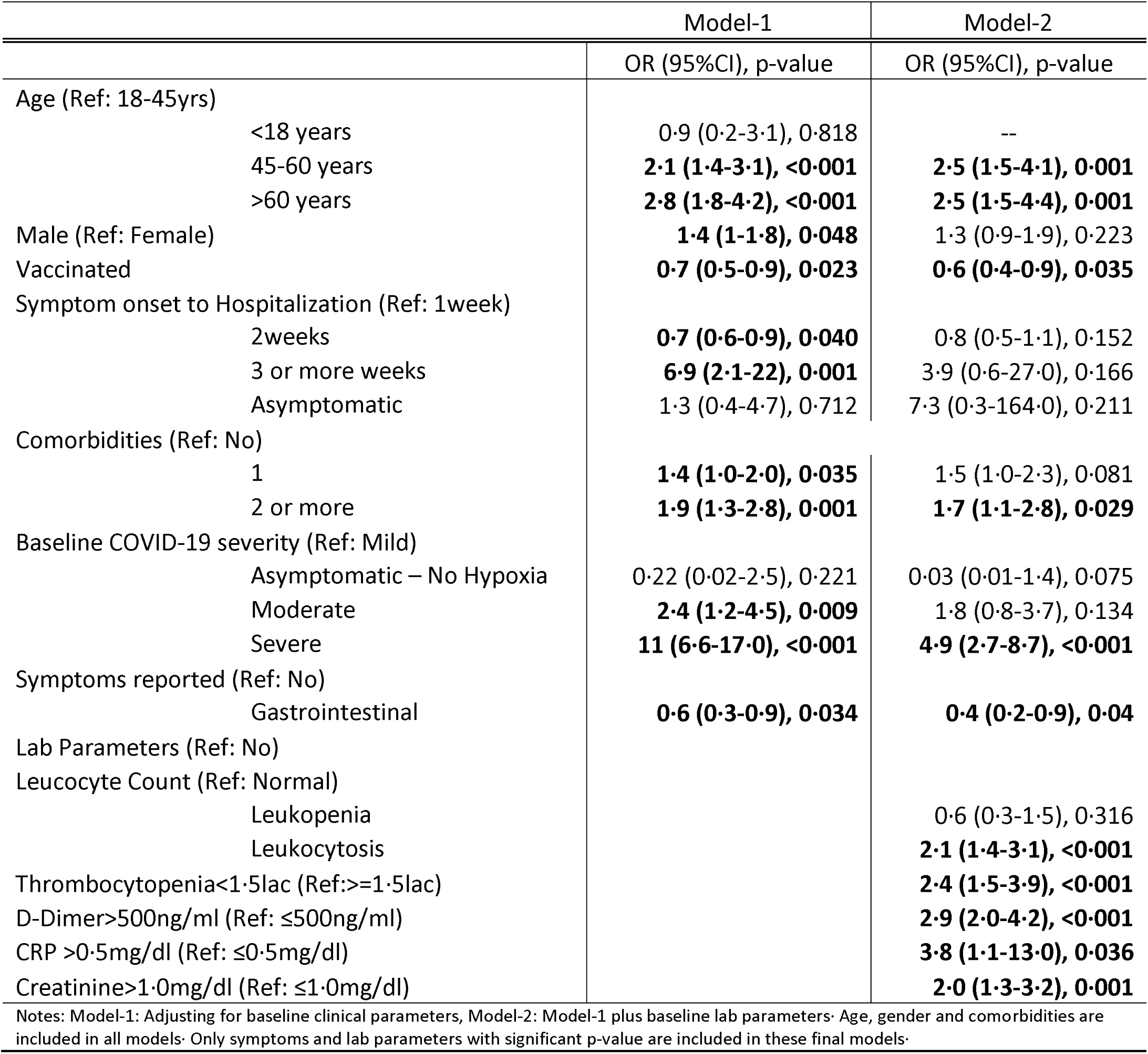
Logistic regression model for developing Critical Illness

**Supplementary Table 2:** Factors predicting deterioration during hospital stay amongst those with no hypoxia at Baseline

|  | Model-1 | Model-2 |
| --- | --- | --- |
|  | OR(95%CI), p-value | OR(95%CI), p-value |
| Age (Ref: 18-45years) |  |  |
| <18 years | 0.78 (0.32-1.88), 0.578 | 0.5 (0.1-2.1), 0.365 |
| 45-60 years | **2.1 (1.38-3.19), 0.001** | 1.4 (0.8-2.4), 0.188 |
| >60 years | **2.44 (1.52-3.91), <0.001** | 1.5 (0.8-2.7), 0.213 |
| Male (Ref: Female) | **1.53 (1.07-2.18), 0.02** | 1.1 (0.72-1.8), 0.588 |
| Vaccinated | 0.71 (0.48-1.05), 0.085 | 0.8 (0.49-1.3), 0.317 |
| Comorbidities (Ref: No) |  |  |
| 1 | **1.5 (1.02-2.22), 0.041** | 1.2 (0.7-1.9), 0.553 |
| 2 or more | **2.13 (1.31-3.45), 0.002** | 1.4 (0.8-2.6), 0.258 |
| Baseline COVID-19 severity (Ref: Mild) |  |  |
| Asymptomatic -No hypoxia | **0.36 (0.14-0.94), 0.036** |  |
| Moderate | **2.22 (1.56-3.15), <0.001** |  |
| Symptoms reported (Ref: No) |  |  |
| Dry cough | **1.49 (1.05-2.1), 0.025** |  |
| Gastrointestinal Symptoms | **1.95 (1.28-2.97), 0.002** |  |
| Fever |  | **2.3 (1.3-4), 0.003** |
| Lab Parameters (Ref: No) |  |  |
| Leucocyte Count (Ref: Normal) |  |  |
| Leukopenia |  | **0.5 (0.3-0.9), 0.026** |
| Leukocytosis |  | **2.3 (1.3-4.0), 0.003** |
| D-Dimer>500ng/ml (Ref: ≤500ng/ml) |  | **1.7 (1.0-3.0), 0.047** |
| CRP >0.5mg/dl (Ref: ≤0.5mg/dl) |  | **9.4 (3.7-24.0), <0.001** |
| Creatinine>1.0mg/dl (Ref: ≤1.0mg/dl) |  | **2.2 (1.2-3.9), 0.010** |
| Notes: Model-1: Adjusting for baseline clinical parameters, Model-2: Model-1 plus baseline lab parameters. Age, gender and comorbidities are included in all the models. Only symptoms and lab parameters with significant p-value are included in these final models. |  |  |

## Notes

Conflicts of interest: None of the authors report any conflict of interest

### Competing Interest Statement

The authors have declared no competing interest.

### Funding Statement

No funding received

### Author Declarations

Institute Ethics committee, All India Institute of Medical Sciences, New Delhi

